# Using Simulation-Based Experiential Learning to Increase Students’ Ability to Analyze Increasingly Complex Global Health Challenges: A Mixed Methods Study

**DOI:** 10.1101/2023.12.19.23300262

**Authors:** Ahmad Firas Khalid, Megan A. George, Clarissa Eggen, Aaranee Sritharan, Faiza Wali, A. M. Viens

**Author notes:** Corresponding author: Ahmad Firas Khalid Address: School of Global Health, Victor Dahdaleh Building, Room 5022, 88 The Pond Road, Toronto, ON, M3J 2S5, Canada. **Contributions to the Literature** This study contributes to the burgeoning literature on experiential learning in health sciences, particularly in the context of global health governance. It employs a robust mixed-method design to provide both breadth and depth of insights into the effectiveness of a simulation-based learning platform. This could be a blueprint for the development of future simulation-based educational initiatives in global health and related fields.

## Abstract

Traditional didactic teaching approaches fall short of adequately supporting diverse student learning styles. Complementing didactic teaching approaches with simulation-based experiential learning has shown promise in bridging the gap between theoretical knowledge and practical application. However, few studies have rigorously examined the outcomes of this approach in global health education and training. This study aims to evaluate the impact of the World Health Organization World Health Assembly Simulation (WHA Sim), a simulation-based experiential learning platform designed to enhance the application of students’ knowledge and skills in practical settings relating to global health governance.

We employed a sequential mixed-method study between September 2022 and July 2023, starting with an anonymous survey among undergraduate students in the Faculty of Health at York University, in order to evaluate self-reported quantitative metrics related to students’ understanding of simulation-based learning prior to the WHA Sim. We also conducted qualitative interviews among participants of the WHA Sim from diverse health disciplines, aiming to capture multi-disciplinary perspectives. Data was analyzed using simple descriptive statistics for the quantitative data and a framework analysis for qualitative data.

Among 39 survey respondents, 18 were interviewed. The WHA Sim bolstered a wide array of skills, including research capabilities, critical analysis, time management, and organizational effectiveness. Participants also reported improvements in effective interpersonal communication, public speaking, networking, and solution-driven dialogues. Interpersonal skills like collaboration and leadership were notably improved. Moreover, the simulation provided an enhanced understanding of complex issues and offered a fertile ground for career preparation, filling existing knowledge gaps more effectively than traditional learning environments.

Findings demonstrate the value of simulation-based experiential learning among undergraduate students, which illustrates how the WHA Sim can serve as an effective learning tool within global health education and training.

## Introduction

The escalating frequency of health and humanitarian crises has significantly fueled students’ engagement with global health topics and institutions, such as the World Health Organization, responsible for addressing these intricate challenges (1–4). Amidst this heightened interest, exacerbated by events like the COVID-19 pandemic, simulation-based experiential learning stands out as a transformative educational approach. It offers students a tangible application of theoretical knowledge and practical skills to real-world dilemmas; a method proven to bridge the gap between classroom theory and applied execution. However, a knowledge gap exists in how to effectively develop and implement these simulations to enhance students’ analysis and problem-solving skills concerning complex global health issues. Previous studies vouching for experiential learning’s efficacy often lack rigorous validation, leaving room for enhancement in this educational approach. Moreover, the deployment of simulations, especially in the realm of public and global health education, is a relatively untapped but burgeoning strategy.

Addressing this gap, our study introduces a novel approach by utilizing a simulation of the World Health Organization’s World Health Assembly (WHA), replicating its role as the paramount decision-making body in global health. The WHA Simulation (WHA Sim) seeks to enhance knowledge and skills around collaborative governance approaches involving multi-sectoral and multi-jurisdictional global challenges, such as those found in the Sustainable Development Goals (SDGs). It enables the generation and testing of innovative and feasible solutions on the simulation theme in the form of research briefs, debates, and formal resolutions. This endeavor not only promises to augment the educational toolkit with a validated, immersive experience but also prepares students more effectively for real-world challenges, thereby enhancing the overall quality of global health education and training activities.

According to classic experiential learning theory, individuals gain knowledge through concrete experience and abstract conceptualization and through reflective observation and active experimental (5, 6). In other words, the experience of engaging in the learning activity leads to new knowledge, learning, and skills development. Simulation-based learning in global health represents an innovative approach to experiential education, designed to extend beyond traditional classroom boundaries. This method facilitates the exploration and development of practical, effective strategies to address the multifaceted challenges inherent in global health. It also allows students to be better prepared with the types of skills required in the real-world job market because they will be working on developing their writing skills (e.g., policy briefs), communication skills (e.g., effectively presenting key points), negotiation skills (e.g., identifying areas of agreement/disagreement and how to manage them), leadership skills (e.g., managing tight deadlines to deliver on resolutions), and team-building skills (e.g., working closely with other delegates on arriving at consensus) (3).

This study has three primary objectives. First, to detail the creation and implementation of the WHA Sim, thoroughly assessing its effectiveness as a teaching tool in undergraduate global health programs or initiatives. Second, to collect firsthand student insights on how simulation-based experiential learning enhances their skills in analyzing complex issues and formulating innovative solutions. Third, to expand our understanding of how this type of experiential learning in a global health simulation can better prepare students for future leadership positions in global health.

## Methods

### Design

We used a sequential mixed-method study design. This type of design combines elements of qualitative and quantitative research approaches to ensure that we arrive at the breadth and depth of understanding of the extent to which simulation-based experiential learning increases students’ ability to analyze increasingly complex problems and develop innovative solutions. First, we conducted general and targeted needs assessments through an anonymous survey of students enrolled in the Faculty of Health at York University in Toronto, Canada. The survey focused on identifying the views and experiences of students’ engagement in simulation-based experiential learning. Second, we used the insights generated from the survey to help inform qualitative interviews with students who took part in the WHA Sim week to derive an in-depth understanding of the knowledge they gained from the experience and feedback on ways to improve the delivery of simulation-based experiential learning to students. Findings were supported by observations of PI (Khalid, AF) at the WHA Sim.

### Characteristics of participants

We purposively sampled current undergraduate students completing their studies in the Faculty of Health. We aimed to survey and interview a diverse group of students at the undergraduate level in programs/faculties of basic science, global health, nursing, health policy and management. The diversity in students is designed to depict multidisciplinary participation similar to real-world global health work and to encourage interprofessional education.

### Participant recruitment and sample size

A two-stage sampling approach was used to identify and recruit key informants (7, 8). The first stage included recruiting students from various disciplines in the Faculty of Health. To recruit these participants, we created a York University centrally supported learning management system (eClass) announcement distributed to Faculty of Health members to share on their course eClass site. We also sent out emails to Faculty of Health Program Administrators to share the advertisement of the study with students via email. Finally, the research team created a video to invite students to participate in the study, which was shared on the Faculty of Health Twitter page and on public domain profiles. The link to the survey was sent to the interested students who would contact us after checking their eligibility and getting their consent. To incentivize students’ participation in our survey, we offered them a raffle for an Amazon gift card. The second stage involved respondent-driven sampling by which students in the first stage were asked to identify any additional informants. We planned to sub-sample participants for the semi-structured interviews from the pool of students who responded to the survey and enrolled in the WHA Sim. Given that our goal is to gain information from limitedly available and ‘convenient’ students to access, we anticipated a sample size of 35 – 50 students. We recruited 39 participants from our first stage of sampling, and 18 additional participants were identified through snowball sampling.

## Data collection methods

### Phase one: quantitative study

Participants completed an online self-assessment survey administered using Microsoft Office 360 Forms (survey instrument available as an e-appendix). The survey was in English and consisted of three parts: (1) questions focusing on participants’ background and demographic characteristics, (2) questions that captured participants’ comfort and confidence in key practice skills, and (3) questions around participants’ experiences and preferences with simulation-based learning.

### Phase two: qualitative study

Interviews with students who responded to the survey and participated in the WHA Sim were conducted in the weeks following WHA Sim week to encourage a high completion rate. Students were invited to engage in a 30-minute Zoom interview to share their perspectives on the WHA Sim experience. The discussion focused on several aspects: enhancing the WHA Sim platform, such as adjusting the simulation’s pace and timing; identifying the key skills and competencies gained through participation; evaluating their readiness to assume leadership roles in addressing global health issues; and considering the educational value of the simulation as an integral part of their learning experience. Students were also asked to reflect on their main takeaway from participating in simulation-based learning using open-ended questions.

The interviews were audio-recorded after receiving permission from the participants. Audio recordings were transcribed verbatim (MG, CE, AS, FW) and the written transcriptions were used for data analysis. Potentially identifying information (e.g., name) was removed at the time of transcription. The language of the interview was in English. A pilot interview with a research team member, which was not included in the actual study, was conducted to refine the interview questions.

### Data analysis

A mixed methods approach to data analysis was employed. For the survey, quantitative data was summarized using simple descriptive statistics (numbers, percentages, frequencies, and cross-tabulation). Data analysis included calculating descriptive statistics related to all assessed measures, including the background of participants, their experiences with experiential learning and their attitudes towards participating in simulation-based learning. For the semi-structured interview data, we used a deductive framework analysis approach for our collected data (9, 10). Framework analysis is a qualitative method that can be applied to research that has specific questions and a limited time frame (9). This approach allowed us to describe and interpret what is happening in a particular setting (i.e., engaging in simulation-based learning) (10). It involved a five-step process: familiarization (i.e., immersing ourselves in collected data making notes of key ideas and recurrent themes), identifying a thematic framework (i.e., recognizing emerging themes), indexing (i.e., using NVivo to identify sections of data that correspond to particular themes), charting (i.e., arranging identified sections of data into table exhibits), and mapping and interpretation (i.e., analyzing key characteristics from the exhibits) (10).

## Results

We first outline the detailed planning and execution process of the WHA Sim, followed by an overview of our participants. Subsequently, we present an analysis of how global health simulation-based experiential learning contributes to preparing students for future leadership roles in global health. We highlight the key themes that emerged, demonstrating the enhanced skills students developed through their engagement in the WHA Sim.

### WHA Sim

The development of the WHA Sim began with the critical choice of a relevant global health theme. For the 2023 iteration, “Building Global Solidarity for Worldwide Health Security” was selected, spotlighting urgent health emergencies, especially pandemics, within both health and humanitarian domains. This theme was particularly pertinent given recent global health crises, like COVID-19 and Ebola, underscoring the dire need for enhanced crisis readiness and response systems. It also aligned with previous endeavors, such as the 2021 Model WHO Simulation Week in London, which similarly focused on pandemic readiness and recovery, incorporating elements like the One Health strategy, healthcare access, misinformation control, and safeguarding the well-being of healthcare workers. In line with our commitment to inclusivity and accessibility, we did not charge students to participate in the simulation. This decision was crucial to ensure maximum participation and access.

The first day of the WHA Sim was delivered virtually, aimed at equipping delegates with essential skills such as understanding WHA Sim protocols, crafting position papers, and mastering debate strategies. Prior to the simulation, students were engaged in the critical task of drafting position papers. This activity necessitated thorough research and teamwork, leading students to formulate astute policy suggestions and innovative approaches. Such early involvement fostered a sense of accountability and bolstered students’ confidence and active participation in subsequent discussions.

The following day transitioned to in-person simulation activities at York University. It featured an inaugural ceremony and an insightful panel discussion with global health luminaries, offering extensive perspectives on the chosen theme. After a short social break, participants plunged into the Opening Plenary and diversified Committee Sessions, addressing key issues like Public Health Emergencies, Infodemic Management, and Universal Health Coverage. These sessions were carefully organized by fellow students who had been appointed to roles within the Delegate Advisory and Information System (DIAS) well before the simulation began. The DIAS team included positions such as Chair, Vice-chair, and Secretariat. These students played crucial roles in creating a realistic World Health Assembly simulation experience. Additionally, they were instrumental in coordinating the drafting of position papers, as mentioned in the previous paragraph, ensuring a structured and authentic simulation process. Their leadership, marked by skilled diplomacy and an exceptional knack for consensus-building, was a standout aspect of the simulation. Notably, each committee was enriched by the presence of a technical expert, an esteemed global health scholar, who deepened the debates with valuable insights and research-based knowledge.

The final day focused on the joint effort of creating and ratifying resolutions, highlighting the importance of collaborative decision-making. The resolutions and materials produced by participants were preserved for future sharing with experts at the World Health Organization. After a global health career panel and additional plenary discussions, the event concluded with parting words from notable figures like Dr. Tedros Ghebreyesus and Dr. Peter Singer of the WHO. This capstone day was not just about dialogue but also involved vital exchanges of inventive concepts and cooperative problem-solving – all crucial for fortifying global health governance. The event culminated in an awards ceremony, recognizing and celebrating the remarkable efforts and contributions of participants, particularly those who excelled in position paper submissions in each committee.

Figure 1. The four components that make up a WHA Simulation — role-based participant interaction, debate, discussion alliance formation, and draft resolution process.

**Figure 1.**
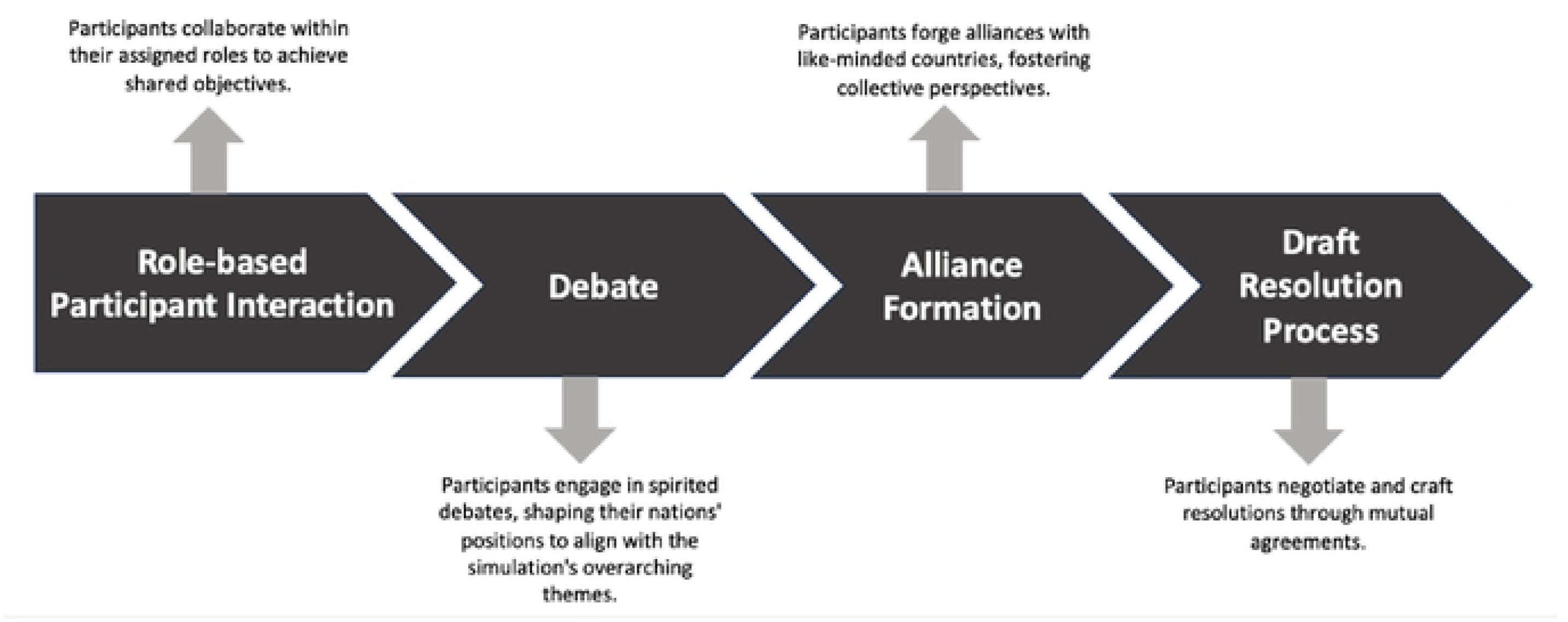
The four components that make up a WHA Simulation.

### Participants’ characteristics

There were 39 students who completed the survey and 18 students who completed the semi-structured interviews. Of the 18 participants participating in semi-structured interviews, one participant did not complete the pre-simulation survey. Table 1 presents the characteristics of the participants.

**Table 1.** Characteristics of Research Participants.

| Study Design | # of Participants (n) | Organizational Affiliations | Program of Study | Academic Year | Sex |
| --- | --- | --- | --- | --- | --- |
| Survey | (n=39) | York University (n= 39) | Global Health (n=34) | Fourth-year (n= 21) | 34 F<br>5 M |
|  |  |  | Health Management (n=3) | Third-year (n= 6) |  |
|  |  |  | Biology (n=1) | Second-year (n= 3) |  |
|  |  |  | Biomedical Science (n=1) | First-year (n= 3) |  |
|  |  |  |  | Other (n= 6) |  |
| **Semi-structured interviews** | (n=18) | York University (n= 18) | Global Health (n=17) | Fourth-year (n=11) | 14 F<br>4 M |
|  |  |  | Health Management (n = 1) | Third-year (n=3) |  |
|  |  |  |  | Second-year (n= 1) |  |
|  |  |  |  | Other (n=3) |  |

### Phase 1: Quantitative Results

#### Views and experiences in simulation-based experiential learning

Participants’ experiences and perceptions of simulation-based experiential learning were explored in the study, as presented in Table 2. Regarding prior involvement in simulation-based learning, only 33.33% of students had engaged in such activities, while a majority of 51.28% had no previous involvement, and 15.38% were uncertain. Awareness levels of simulation-based experiential learning were higher, with 66.67% of students knowledgeable about this educational approach, 17.95% unaware, and 15.38% only somewhat aware. Familiarity with the World Health Organization’s World Health Assembly was prevalent, with 76.92% having heard of it, though only 17.95% were knowledgeable about its purpose, and 5.13% had never encountered it. The belief in the necessity of simulation-based learning for their education was strong among students, with 59.97% in agreement, though a notable 38.46% remained unsure, and a minimal 2.56% disagreed. Inquiry into the integration of any form of simulation-based learning during their degree revealed mixed responses: 38.46% confirmed its presence, 41.03% denied it, and 20.51% were uncertain. Assessing the efficacy of simulation-based learning in preparing students for real-world experiences yielded positive views: 48.72% found it very effective and 51.28% deemed it somewhat effective, with none of the respondents labeling it as ineffective in any degree. These results suggest a recognition of the value of simulation-based learning among students, although exposure to and comprehensive understanding of such educational strategies appear to be varied.

**Table 2.**
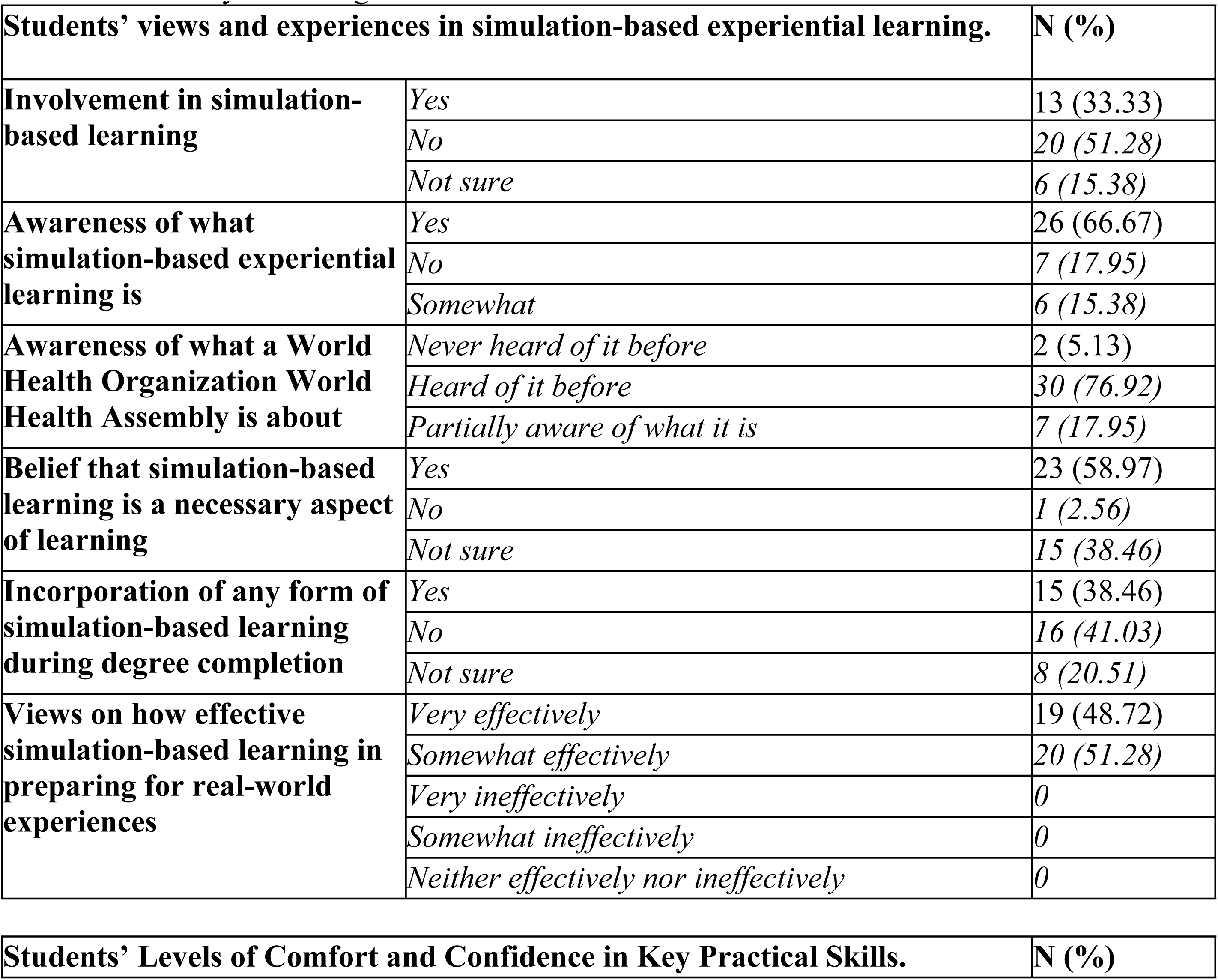
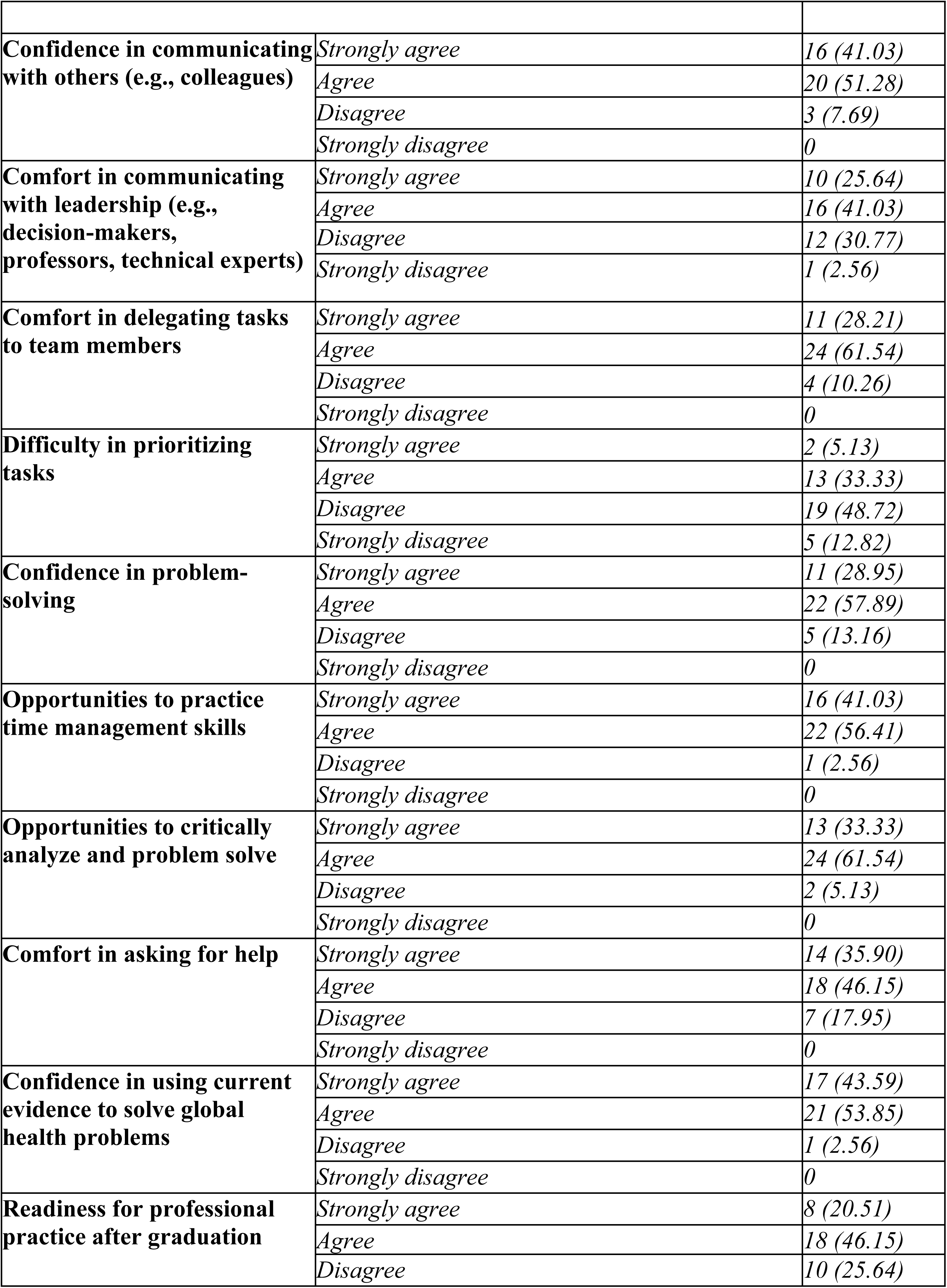
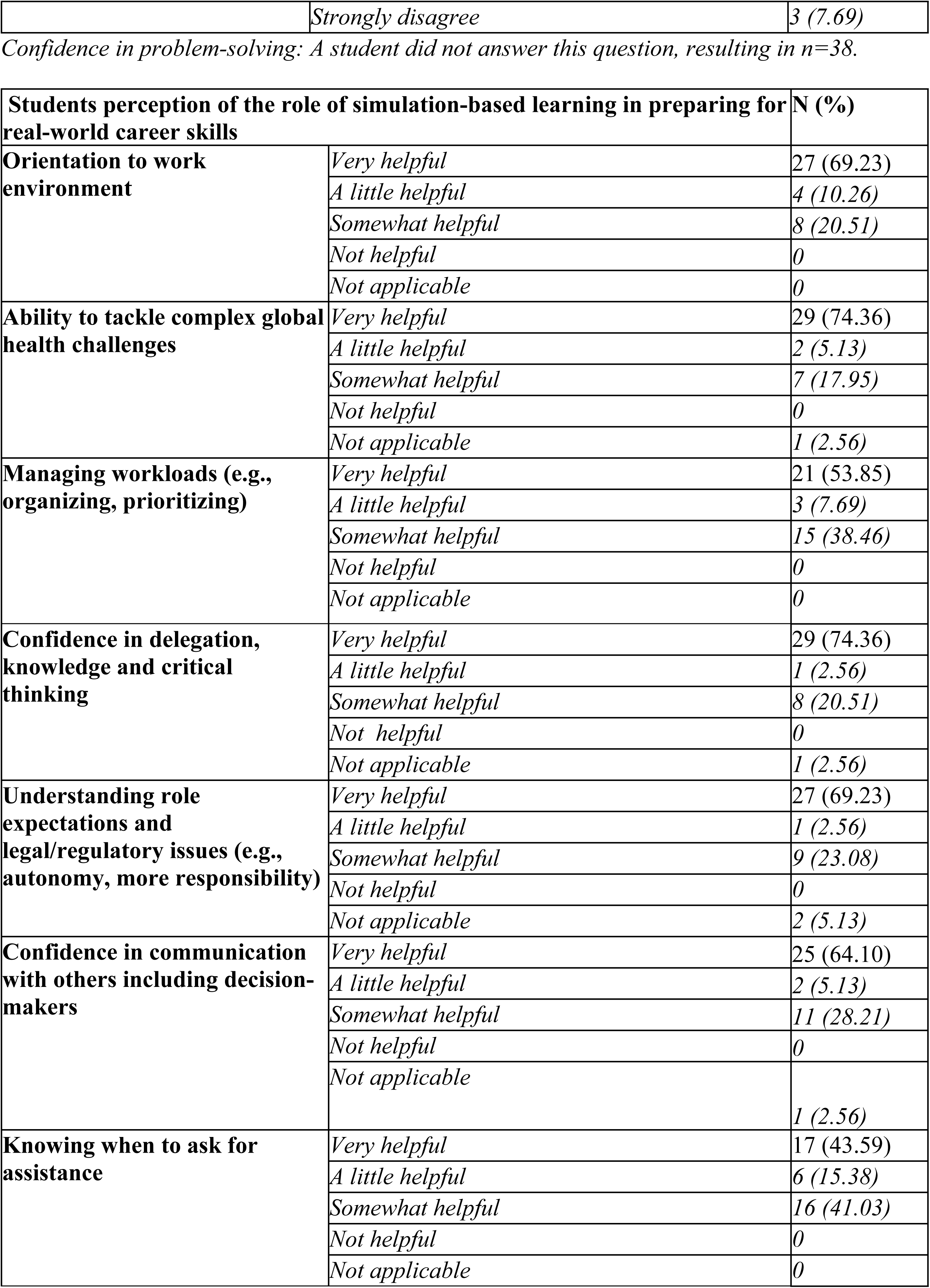
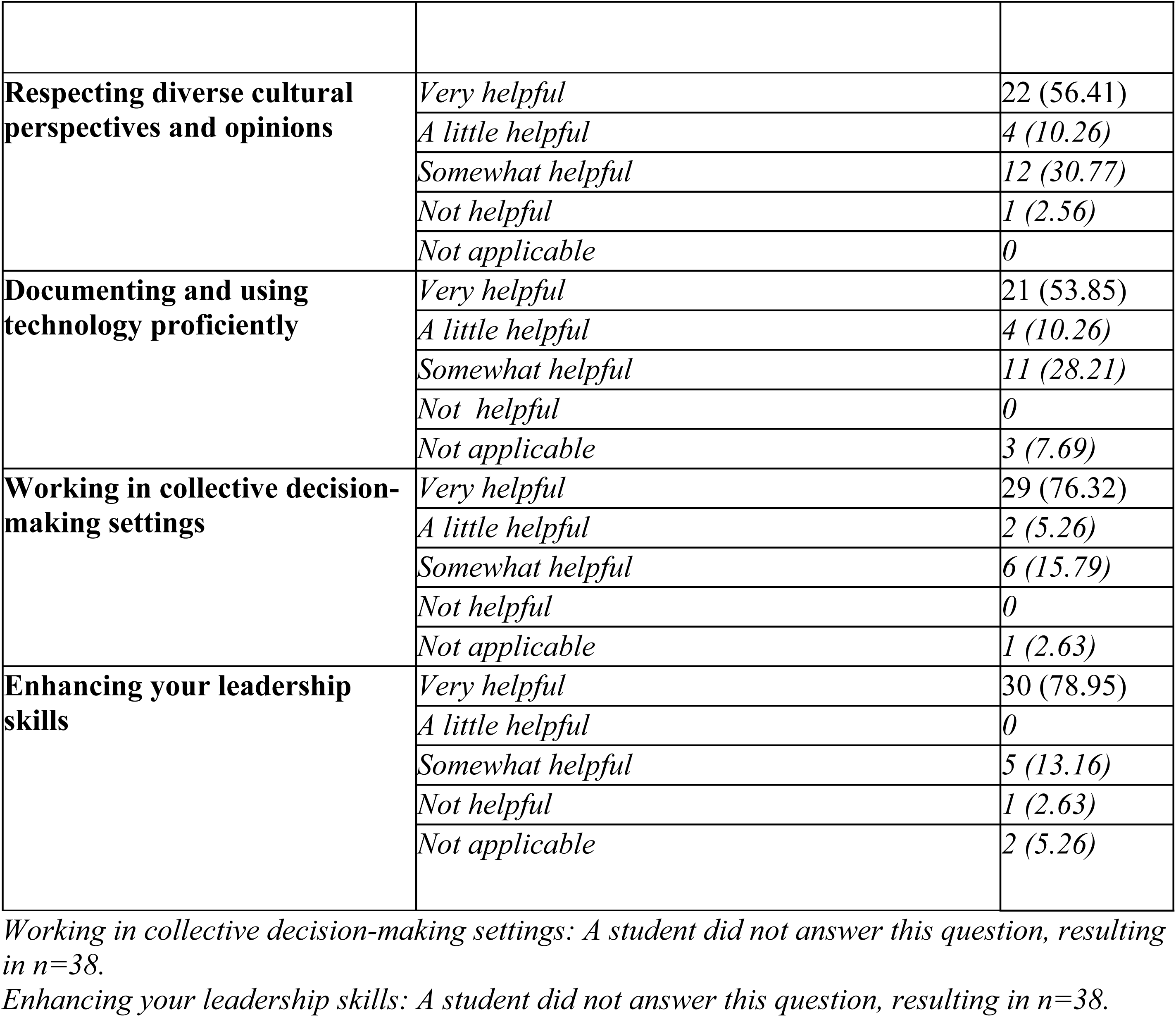
Summary of findings table.

| **Students' views and experiences in simulation-based experiential learning.** |  | **N (%)** |
| --- | --- | --- |
| **Involvement in simulation-based learning** | *Yes* | 13 (33.33) |
|  | *No* | 20 (51.28) |
|  | *Not sure* | 6 (15.38) |
| **Awareness of what simulation-based experiential learning is** | *Yes* | 26 (66.67) |
|  | *No* | 7 (17.95) |
|  | *Somewhat* | 6 (15.38) |
| **Awareness of what a World Health Organization World Health Assembly is about** | *Never heard of it before* | 2 (5.13) |
|  | *Heard of it before* | 30 (76.92) |
|  | *Partially aware of what it is* | 7 (17.95) |
| **Belief that simulation-based learning is a necessary aspect of learning** | *Yes* | 23 (58.97) |
|  | *No* | 1 (2.56) |
|  | *Not sure* | 15 (38.46) |
| **Incorporation of any form of simulation-based learning during degree completion** | *Yes* | 15 (38.46) |
|  | *No* | 16 (41.03) |
|  | *Not sure* | 8 (20.51) |
| **Views on how effective simulation-based learning in preparing for real-world experiences** | *Very effectively* | 19 (48.72) |
|  | *Somewhat effectively* | 20 (51.28) |
|  | *Very ineffectively* | 0 |
|  | *Somewhat ineffectively* | 0 |
|  | *Neither effectively nor ineffectively* | 0 |

| **Students' Levels of Comfort and Confidence in Key Practical Skills.** | **N (%)** |
| --- | --- |
| **Confidence in communicating with others (e.g., colleagues)** | *Strongly agree* | 16 (41.03) |
|  | *Agree* | 20 (51.28) |
|  | *Disagree* | 3 (7.69) |
|  | *Strongly disagree* | 0 |
| **Comfort in communicating with leadership (e.g., decision-makers, professors, technical experts)** | *Strongly agree* | 10 (25.64) |
|  | *Agree* | 16 (41.03) |
|  | *Disagree* | 12 (30.77) |
|  | *Strongly disagree* | 1 (2.56) |
| **Comfort in delegating tasks to team members** | *Strongly agree* | 11 (28.21) |
|  | *Agree* | 24 (61.54) |
|  | *Disagree* | 4 (10.26) |
|  | *Strongly disagree* | 0 |
| **Difficulty in prioritizing tasks** | *Strongly agree* | 2 (5.13) |
|  | *Agree* | 13 (33.33) |
|  | *Disagree* | 19 (48.72) |
|  | *Strongly disagree* | 5 (12.82) |
| **Confidence in problem-solving** | *Strongly agree* | 11 (28.95) |
|  | *Agree* | 22 (57.89) |
|  | *Disagree* | 5 (13.16) |
|  | *Strongly disagree* | 0 |
| **Opportunities to practice time management skills** | *Strongly agree* | 16 (41.03) |
|  | *Agree* | 22 (56.41) |
|  | *Disagree* | 1 (2.56) |
|  | *Strongly disagree* | 0 |
| **Opportunities to critically analyze and problem solve** | *Strongly agree* | 13 (33.33) |
|  | *Agree* | 24 (61.54) |
|  | *Disagree* | 2 (5.13) |
|  | *Strongly disagree* | 0 |
| **Comfort in asking for help** | *Strongly agree* | 14 (35.90) |
|  | *Agree* | 18 (46.15) |
|  | *Disagree* | 7 (17.95) |
|  | *Strongly disagree* | 0 |
| **Confidence in using current evidence to solve global health problems** | *Strongly agree* | 17 (43.59) |
|  | *Agree* | 21 (53.85) |
|  | *Disagree* | 1 (2.56) |
|  | *Strongly disagree* | 0 |
| **Readiness for professional practice after graduation** | *Strongly agree* | 8 (20.51) |
|  | *Agree* | 18 (46.15) |
|  | *Disagree* | 10 (25.64) |
|  | *Strongly disagree* | 3 (7.69) |

Confidence in problem-solving: A student did not answer this question, resulting in n=38.
| **Students perception of the role of simulation-based learning in preparing for real-world career skills** |  | **N (%)** |
| --- | --- | --- |
| **Orientation to work environment** | *Very helpful* | 27 (69.23) |
|  | *A little helpful* | 4 (10.26) |
|  | *Somewhat helpful* | 8 (20.51) |
|  | *Not helpful* | 0 |
|  | *Not applicable* | 0 |
| **Ability to tackle complex global health challenges** | *Very helpful* | 29 (74.36) |
|  | *A little helpful* | 2 (5.13) |
|  | *Somewhat helpful* | 7 (17.95) |
|  | *Not helpful* | 0 |
|  | *Not applicable* | 1 (2.56) |
| **Managing workloads (e.g., organizing, prioritizing)** | *Very helpful* | 21 (53.85) |
|  | *A little helpful* | 3 (7.69) |
|  | *Somewhat helpful* | 15 (38.46) |
|  | *Not helpful* | 0 |
|  | *Not applicable* | 0 |
| **Confidence in delegation, knowledge and critical thinking** | *Very helpful* | 29 (74.36) |
|  | *A little helpful* | 1 (2.56) |
|  | *Somewhat helpful* | 8 (20.51) |
|  | *Not helpful* | 0 |
|  | *Not applicable* | 1 (2.56) |
| **Understanding role expectations and legal/regulatory issues (e.g., autonomy, more responsibility)** | *Very helpful* | 27 (69.23) |
|  | *A little helpful* | 1 (2.56) |
|  | *Somewhat helpful* | 9 (23.08) |
|  | *Not helpful* | 0 |
|  | *Not applicable* | 2 (5.13) |
| **Confidence in communication with others including decision-makers** | *Very helpful* | 25 (64.10) |
|  | *A little helpful* | 2 (5.13) |
|  | *Somewhat helpful* | 11 (28.21) |
|  | *Not helpful* | 0 |
|  | *Not applicable* | 1 (2.56) |
| **Knowing when to ask for assistance** | *Very helpful* | 17 (43.59) |
|  | *A little helpful* | 6 (15.38) |
|  | *Somewhat helpful* | 16 (41.03) |
|  | *Not helpful* | 0 |
|  | *Not applicable* | 0 |
| **Respecting diverse cultural perspectives and opinions** | *Very helpful* | 22 (56.41) |
|  | *A little helpful* | 4 (10.26) |
|  | *Somewhat helpful* | 12 (30.77) |
|  | *Not helpful* | 1 (2.56) |
|  | *Not applicable* | 0 |
| **Documenting and using technology proficiently** | *Very helpful* | 21 (53.85) |
|  | *A little helpful* | 4 (10.26) |
|  | *Somewhat helpful* | 11 (28.21) |
|  | *Not helpful* | 0 |
|  | *Not applicable* | 3 (7.69) |
| **Working in collective decision-making settings** | *Very helpful* | 29 (76.32) |
|  | *A little helpful* | 2 (5.26) |
|  | *Somewhat helpful* | 6 (15.79) |
|  | *Not helpful* | 0 |
|  | *Not applicable* | 1 (2.63) |
| **Enhancing your leadership skills** | *Very helpful* | 30 (78.95) |
|  | *A little helpful* | 0 |
|  | *Somewhat helpful* | 5 (13.16) |
|  | *Not helpful* | 1 (2.63) |
|  | *Not applicable* | 2 (5.26) |

*Working in collective decision-making settings: A student did not answer this question, resulting in n=38.*
*Enhancing your leadership skills: A student did not answer this question, resulting in n=38.*

#### Comfort and confidence in key practical skills

Upon completing an online self-assessment survey, participants revealed diverse perceptions of their practical skills and preparedness for professional practice, as detailed in Table 2. A substantial majority expressed high confidence in their communication abilities (92.31% total for strongly agree and agree) and problem-solving skills (86.84% total for strongly agree and agree). However, comfort in interacting with leadership was less robust, with only 66.67% in the affirmative and a not insignificant 33.33% expressing discomfort. Delegating tasks was viewed favorably by 89.75%, yet participants acknowledged challenges in task prioritization, with a majority (61.54%) experiencing difficulties. The survey also indicated strong recognition of opportunities to hone time management skills (97.44%) and to engage in critical problem-solving (94.87%). Most participants were comfortable seeking help (81.05%), and nearly all expressed confidence in utilizing evidence-based approaches to global health issues (97.44%). In contrast, readiness for professional practice post-graduation elicited a more measured response, with a combined affirmative response of only 66.66%, suggesting a gap in perceived career preparedness. These insights, derived from the self-assessment, emphasize the need for educational programs to strategically reinforce areas of confidence while addressing identified gaps in professional readiness.

#### Preparing for real-world career skills

In assessing the impact of simulation-based learning on real-world career skill preparation (Table 3), students predominantly acknowledged its substantial benefits across multiple competencies. A majority found it “very helpful” in familiarizing themselves with work environments (69.23%), tackling global health challenges (74.36%), and enhancing critical skills such as delegation, knowledge application, and critical thinking (74.36%). Similarly, understanding role expectations and legal matters (69.23%), effective communication (64.10%), and cultural respect (56.41%) were areas where simulations were considered significantly advantageous. Though the perceived helpfulness slightly diminished in aspects like managing workloads (53.85%), technology use (53.85%), and recognizing when to seek help (43.59%), the majority still recognized the value. Impressively, collective decision-making (76.32%) and leadership skills (78.95%) were areas where simulations were deemed most beneficial. These findings underscore the critical role of simulation-based learning in bolstering practical career skills among students, despite some variability in perceived efficacy across different domains.

**Table 3.** Overview of Student Evaluations on the Influence of Simulation-Based Learning in Enhancing Real-World Career Skills Across Three Core Competency Areas.

| Professional Development | Interpersonal Skills | Knowledge and Understanding |
| --- | --- | --- |
| • Communication<br>• Confidence<br>• Preparation for future career<br>• Leadership | • Teamwork<br>• Critical thinking<br>• Collaboration<br>• Compromise<br>• Organization | • Enhanced understanding of processes<br>• Exposure to complex problems<br>• Making intellectual inquiries<br>• Highlighting knowledge gaps<br>• Retention through applied learning |

### Phase 2: Qualitative results

#### Deepening of knowledge and practical understanding

The analysis of 18 semi-structured interviews reveals a consensus among students: participation in the WHA Sim significantly deepened their practical understanding and knowledge. This finding is crucial in evaluating our research question about the role of simulation-based experiential learning in enhancing students’ problem-solving and innovative thinking in complex global health scenarios.

Students reported themes of enhanced processes understanding, classroom concepts solidification, complex problem exposure, knowledge gaps identification, and concept retention via applied learning. One remarked:

> *“Interacting with various fields and applying knowledge in real-world contexts made me realize my extensive learning over four years… I’ve become a specialist with substantial expertise, which is key for career success.”*

Another highlighted the bridge between theory and practice: *“[…] with the simulation, you can apply theories and see what works and what doesn’t, which enhances learning.”* Participation also improved intellectual engagement. One student shared, “*Being around people who ask questions and engage, I learned to do the same… asking the right questions can build valuable mentor relationships.”*

Furthermore, the WHA Sim encouraged more open, creative problem-solving. A participant noted: *“The absence of a rubric allowed for creative, collaborative solutions, free from strict guidelines.”* Another appreciated the reduced pressure, adding, *“Not being evaluated made it easier to engage and apply my knowledge.”*

These reflections affirm the simulation’s effectiveness in fostering both deep understanding and practical skills essential for addressing global health complexities, thus providing profound insights into our research inquiry.

#### Interpersonal Skills and Collaboration

Many students reported a marked improvement in their ability to work effectively within teams and engage in compromise, essential skills for both problem-solving and leadership. One participant shared,

> *“Collaborating with people who may lack certain background knowledge taught me how to balance giving feedback without coming across as critical or taking control of the project. It’s about expressing ideas clearly without hurting anyone’s feelings and ensuring the final product is something we’re all proud of.”*

Additionally, the WHA Sim seemed to significantly bolster students’ critical thinking skills, a crucial aspect of analyzing complex issues and innovating solutions. One student reflected, “*The simulation made me think out of the box when I’m creating solutions for a certain problem*.” Another noted the complexity of global health challenges: “*We have to think critically about how some of these global health problems actually play out. It’s not a sugar-coated thing where we can just say, oh, let’s just do this, it’s a lot more complex*.”

Furthermore, the simulation underscored the power of collective problem-solving and fostered a solution-driven environment. A student observed, “*It was an opportunity for me to work with my classmates, which is something outside of the classroom that we don’t often get a chance to do.*” This sentiment was echoed by another:

> *“Working in smaller groups allowed us to brainstorm ideas collaboratively, resulting in more innovative solutions. Someone would suggest an idea, and others would build upon it, leading to the evolution and improvement of our concepts over time.”*

The experiential learning setup of the WHA Sim also seemed to inspire a deeper understanding of the practicalities and challenges of real-world global health efforts. Students noted insights gained into the intricacies of change management and the patience required for impacting large-scale perspectives. One stated, “*I think that’s one of the things that the WHA Sim helped with is the practicality of how a lot of this work is being done behind the scenes*”, while another reflected on the complexity of convincing diverse stakeholders, “*It’s about like changing entire populations’ perspectives, which requires a lot more patience and intentionality than trying to convince one person at a time.*”

Moreover, the simulation seemed to act as a catalyst for students to visualize themselves as change agents, particularly regarding the Sustainable Development Goals (SDGs). One student felt the experience was instrumental in inspiring new ideas and potential research directions, asserting,

> *“Having that simulation will help inspire new ideas… we can help be the agents of change that help facilitate issues, especially the SDG goals. We can really help resolve that and find ways to resolve that. I think the simulation is good to have to find solutions for the world’s issues, if that makes sense.”*

#### Professional development

Participants identified the WHA Sim as a unique opportunity for experiential learning that contributed significantly to their professional growth. The immersive nature of the simulation allowed students to “*think on their feet,”* a stark contrast to traditional classroom presentations. This environment also fostered public speaking skills because, as one student articulated, “[…] *you’re building that skill*” in a non-judgmental setting.

Moreover, the WHA Sim provided students with a realistic preview of potential career paths, as well as an environment to assess and develop their competencies. One student contemplated their future, stating,

> *“I think just from like picturing myself working later on. As someone who’s always said they would love to work in the World Health Organization, I was like okay it would be good to participate in this kind of opportunity and just get a sense for what it would feel like and the exact skills that are needed just to see if what I currently have would be enough or what I need to improve on.”*

This reflection underscores the simulation’s role in career preparation and skill assessment, further echoed by another participant: *“[…] that helped me to identify my weaknesses and actually see it not even like just I know, but I’ll be in this part and seeing that I actually need to work on my articulation*”.

Students also recognized the simulation’s efficacy in presenting “*real-world career options*”, giving them the confidence and preparation for their future roles in global health. The practical experience of the WHA Sim, far removed from theoretical learning, offered students insights into the complexities of global health leadership and diplomacy. As one student succinctly put it, it is about the “confidence that it gave me.”

Furthermore, the WHA Sim environment nurtured leadership skills in ways traditional classrooms could not. Participants were compelled to take up active leadership roles, navigating challenges and decision-making without a predefined path. One student reflected on this growth: *“The main thing is how to be a leader. […] But this was new and so you had to step up […]”*.

## Discussion

Our study examined the extent of simulation-based experiential learning in enhancing students’ abilities to analyze complex issues and devise innovative solutions, particularly within the realm of global health. A significant finding was that despite varied exposure to simulation-based experiential learning, there was a unanimous recognition of its value, with students attesting to its effectiveness in bridging theoretical knowledge and practical skills. Notably, the WHA Sim deepened participants’ comprehension of global health complexities, encouraging creative, problem-solving, and fostering intellectual engagement, critical thinking, and effective collaboration. The simulation environment also significantly bolstered participants’ confidence in communication, problem-solving, and leadership, despite some expressing initial discomfort in interacting with leadership and acknowledging challenges in task prioritization.

In terms of the implications for preparing students for future leadership positions in global health, the study highlighted the indispensable role of simulation-based experiential learning in professional development. The WHA Sim was particularly impactful, providing a realistic preview of potential career paths and serving as a platform for students to evaluate and hone their skills, significantly contributing to their career preparedness. Participants reported enhancements in various competencies, including public speaking, critical thinking, and leadership, attributing this growth to the unique challenges presented by the simulation. The immersive experience allowed students to project themselves into their future careers, particularly those aspiring to positions at organizations like the WHO, offering them a practical perspective on the skills they need to develop or refine. Moreover, the study found that the simulation’s practical experience, distinct from theoretical learning, offered invaluable insights into the intricacies of global health leadership and diplomacy, empowering students with greater insight into what it would take to assume a future leadership role.

### Findings in relation to other studies

The skill sets honed by students during the WHA Sim, encompassing writing, communication, leadership, and teamwork, reflect the competencies sought in the contemporary job market. These developed abilities resonate with Kolb’s experiential learning theory, underscoring the necessity to diminish the skill gap between academic learning and professional requirements (5). Notably, the simulation’s real-world fidelity, whether in mirroring the actual WHA procedures or emulating potential workplace challenges and resolutions, was pivotal. This adherence to realism not only enriched the learning experience but also aligned with the National Society for Experiential Education’s best practices (11). The experiential approach’s veracity served as a catalyst for student motivation, with learners perceiving the knowledge gained as both pertinent and transferrable to their prospective careers (12).

Active engagement’s centrality in academic achievement is well-documented, with studies, like Hake (1998) showing that interactive methods in an introductory physics course nearly doubled the conceptual comprehension scores relative to traditional lectures (13). While our study did not evaluate test scores, the WHA Sim’s interactive nature reportedly bolstered knowledge retention and the practical application of concepts among students. Additionally, the origins of simulation-based learning are rooted in fields like aviation and military (14). Its effectiveness is well-established within certain disciplines (15–17); however, these analyses typically concentrate on singular fields, lacking a cross-disciplinary perspective. Our research contributes a multifaceted view, expanding comprehension of simulation-based experiential learning’s effects across various sectors, including healthcare, public health, decision-making, and policy development (4). This multidisciplinary approach offers broader insights into the method’s implications across diverse fields.

### Strengths and limitations

There are three strengths to this study. Firstly, this is the first study to address the knowledge gap in our understanding of the direct impact of simulation-based experiential learning on global health students’ preparedness for real-world challenges and leadership roles. By focusing on both the quantitative self-assessments of students and qualitative feedback obtained through semi-structured interviews, the study offers a comprehensive perspective, balancing statistical data with personal experiences and insights. Secondly, the study’s incorporation of a simulation of the World Health Assembly provides a unique, immersive learning environment that closely mirrors the complexities and dynamics of global health diplomacy and decision-making processes. This realism adds a layer of practical experience to theoretical knowledge, helping to evaluate students’ problem-solving and leadership skills in a manner that more traditional, classroom-based learning methodologies could not replicate.

Thirdly, the diverse range of participants, encompassing various academic backgrounds, cultural perspectives, and levels of expertise, contributes to the robustness of the data and the universality of the findings. This diversity ensures that the insights and conclusions drawn are not overly specific to a particular subgroup of students but are instead indicative of broader trends and potentials within the realm of global health education. One limitation is the potential for self-assessment bias in the quantitative sections of the research, as students might overestimate or underestimate their competencies and experiences, which could affect the reliability of the data. To address this limitation regarding potential self-assessment bias, we designed the survey with a five-point scale for self-evaluation. This approach helped students accurately identify their level of competency, offering a suitable option even for those who considered themselves at an intermediate level.

### Implications for teaching practice

Our research proposes five strategies for enhancing simulation-based experiential learning to strengthen students’ capabilities in dissecting complex issues and crafting innovative solutions in global health, while also fortifying their professional preparedness for careers in this critical sector:

#### 1. Embracing Interdisciplinary Approaches

This involves incorporating diverse academic perspectives and methodologies, encouraging students to think beyond traditional boundaries and understand the multifaceted nature of global health challenges. By engaging with varied disciplines, students gain a holistic view that is essential for effective problem-solving in a complex, interconnected world.

#### 2. Increasing Simulation Realism

By intensifying the authenticity of simulations, including real-world scenarios, stakeholder dynamics, and unexpected challenges, students are immersed in the realities that they will face in professional settings. This heightened realism ensures that learners are tested and adapted to conditions reflecting actual global health crises, enhancing their practical skills and decision-making confidence.

#### 3. Providing Structured Reflection

Post-simulation debriefing sessions with technical experts should be integral, enabling students to reflect on their experiences, actions, and outcomes. Through guided reflection, they can identify strengths, acknowledge weaknesses, and understand the consequences of their decisions, which deepens learning and fosters personal and professional growth.

#### 4. Focusing on Essential Skills

Simulation-based experiential learning should be meticulously designed to cultivate key competencies such as critical thinking, communication, teamwork, and leadership. Tailoring scenarios and roles to target these skills ensures that students are equipped with the essential tools that they will need to be effective leaders and collaborators in the field of global health.

#### 5. Emphasizing Cultural Competency

Given the global nature of public health, it is crucial for simulations to encompass diverse cultural contexts, encouraging students to navigate and respect different cultural perspectives. This exposure prepares them to handle sensitive health issues adeptly, communicate effectively across cultures, and implement solutions that are culturally respectful and sustainable.

The results of our study carry with them some implications for academic institutions, highlighting the necessity to integrate more simulation-based experiential learning into global health curriculums. These findings suggest that simulations, such as the WHA Sim, offers students tangible benefits not always attainable through traditional learning methods or classroom settings. By participating in real-world scenario simulations, students develop critical soft skills, including strategic communication, leadership, problem-solving, and consensus-building, all of which are integral to effective global health leadership and management.

Academic institutions, therefore, should consider expanding their investment in simulation-based experiential learning methodologies. This might entail the development of comprehensive simulation programs, collaboration with global health organizations, or the integration of technology to create immersive, interactive learning environments. Moreover, faculty members could benefit from professional development in simulation facilitation, ensuring that these simulations are as effective and reflective of current global health challenges as possible. Furthermore, the establishment of longitudinal tracking systems to monitor graduates’ career progress post-simulation-based experiential learning could provide valuable data to continuously refine and validate these educational approaches. This commitment not only enhances the educational experience but also ensures that institutions are producing well-equipped, adaptable professionals ready to face the multifaceted challenges present in the global health landscape.

### Future research

Future research should explore the enduring impact of simulation-based experiential learning on the life-long learning and careers of global health students. This involves following the professional development of participants over an extended period to assess how the WHA Sim experience affects their future engagement with training and professional development opportunities, career decisions, leadership roles, and contributions to the global health field. Furthermore, it is important to conduct studies comparing simulation-based experiential learning with alternative educational strategies in global health education. Such studies would clarify the specific advantages and drawbacks of each method, providing valuable insights for shaping curricula and directing educational and training resources.

## Conclusions

In conclusion, the WHA Sim emerges as an invaluable tool in preparing global health students for future leadership positions. It does so by enhancing essential communication skills, enhanced processes understanding, classroom concepts solidification, complex problem exposure, knowledge gaps identification, and concept retention via applied learning offering a realistic appraisal, and understanding of global health careers, and fostering the development of leadership skills in an applied, real-world context.

## Abbreviations

DIAS: Delegate Advisory and Information System
SDG: Sustainable Development Goals
WHA: World Health Assembly
WHA Sim: World Health Assembly Simulation
WHO: World Health Organization

## Declarations

### Ethical approval and consent to participate

Ethical approval was obtained from the Office of Research Ethics (ORE) at York University. Certificate #: e2022-401. We obtained written and verbal consent from all participants.

### Consent for publication

Not applicable.

### Availability of data and material

All data generated or analyzed during this study are included in this published article (and its supplementary information files).

### Authors’ contributions

AFK conceived, analyzed and interpreted the data, and drafted the manuscript. MG, CE, AS, FW assisted in collection of the data. AFK and MG designed the survey and was a major contributor in analyzing and writing quantitative data. MG and AMV were major contributors in interpreting the data and in editing the manuscript. All authors read and approved the final manuscript.

## Data Availability

All data and related metadata underlying the findings reported in the submitted manuscript are part of the submitted article.

## Acknowledgements

The authors extend their gratitude to the team at the Dahdaleh Institute for Global Health Research, the School of Global Health, and the broader York University community for their invaluable support in initiating the first WHA Sim.

